# Improved limit of detection for zoonotic *Plasmodium knowlesi* and *P. cynomolgi* surveillance using reverse transcription for total nucleic acid preserved samples or dried blood spots

**DOI:** 10.1101/2024.04.04.24305339

**Authors:** Kamil A Braima, Kim A Piera, Inke ND Lubis, Rintis Noviyanti, Giri S Rajahram, Pinkan Kariodimedjo, Irbah RA Nainggolan, Ranti Permatasari, Leily Trianty, Ristya Amalia, Sitti Saimah binti Sakam, Angelica F Tan, Timothy William, Jacob AF Westaway, PingChin Lee, Sylvia Daim, Henry Surendra, Nathaniel Christy, Andrew G Letizia, Christopher L Peatey, Mohd Arshil Moideen, Bridget E Barber, Colin J Sutherland, Nicholas M Anstey, Matthew J Grigg

## Abstract

**Background:** Zoonotic *P. knowlesi* and *P. cynomolgi* symptomatic and asymptomatic infections occur across endemic areas of Southeast Asia. Most infections are low-parasitemia, with an unknown proportion below routine microscopy detection thresholds. Molecular surveillance tools optimizing the limit of detection (LOD) would allow more accurate estimates of zoonotic malaria prevalence.

**Methods:** An established ultra-sensitive *Plasmodium* genus quantitative-PCR (qPCR) assay targeting the 18S rRNA gene underwent LOD evaluation with and without reverse transcription (RT) for *P. knowlesi*, *P. cynomolgi* and *P. vivax* using total nucleic acid preserved (DNA/RNA Shield^TM^) isolates and archived dried blood spots (DBS). LODs for selected *P. knowlesi-*specific assays, and reference *P. vivax-* and *P. cynomolgi*-specific assays were determined with RT. Assay specificities were assessed using clinical malaria samples and malaria-negative controls.

**Results:** The use of reverse transcription improved *Plasmodium* species detection by up to 10,000-fold (*Plasmodium* genus), 2759-fold (*P. knowlesi*), 1000-fold (*P. vivax*) and 10-fold (*P. cynomolgi*). The median LOD with RT for the Kamau et al. *Plasmodium* genus RT-qPCR assay was ≤0.0002 parasites/µL for *P. knowlesi* and 0.002 parasites/µL for both *P. cynomolgi* and *P. vivax*. The LODs with RT for *P. knowlesi*-specific PCRs were: Imwong et al. 18S rRNA (0.0007 parasites/µL); Divis et al. real-time 18S rRNA (0.0002 parasites/µL); Lubis et al. hemi-nested *SICAvar* (1.1 parasites/µL) and Lee et al. nested 18S rRNA (11 parasites/µL). The LOD for *P. vivax-* and *P. cynomolgi-*specific assays with RT were 0.02 and 0.20 parasites/µL respectively. For DBS *P. knowlesi* samples the median LOD for the *Plasmodium* genus qPCR with RT was 0.08, and without RT was 19.89 parasites/uL (249-fold change); no LOD improvement was demonstrated in DBS archived beyond 6 years. The *Plasmodium* genus and *P. knowlesi*-assays were 100% specific for *Plasmodium* species and *P. knowlesi* detection, respectively, from 190 clinical infections and 48 healthy controls. Reference *P. vivax-*specific primers demonstrated known cross-reactivity with *P. cynomolgi*.

**Conclusion:** Our findings support the use of an 18S rRNA *Plasmodium* genus qPCR and species-specific nested PCR protocol with RT for highly-sensitive surveillance of zoonotic and human *Plasmodium* species infections.

**Author Summary:** The monkey malaria parasite *Plasmodium knowlesi* has been found to increasingly infect humans across Southeast Asia via the bite of it’s anopheline mosquito vectors. Human infections with a similar monkey parasite, *Plasmodium cynomologi,* have also been reported. The diagnostic tools commonly used to detect these malaria species are often unable to detect very low-level infections. We aimed to to improve surveillance detection tools and blood sample collection methods to detect these zoonotic malaria species and understand the extent of transmission and the burden of disease. This study validated and compared the use of molecular laboratory assays targeting these species. We found that with the use of reverse transcription, large improvements in the limit of detection were possible, by up to 10,000-fold for initial malaria screening, and up to 2759-fold for specific *P. knowlesi* detection. Findings from this study support the use of ultrasensitive detection tools to improve surveillance approaches to emerging zoonotic malaria species.

## BACKGROUND

*Plasmodium knowlesi* is a unicellular protozoan malaria parasite present across Southeast Asia within the geographical range of its natural monkey hosts and vector mosquitoes (1,2). *P. knowlesi* is the most common cause of human malaria in Malaysia; capable of causing severe disease comparable to *P. falciparum* (3–5). Human infections with other genetically similar zoonotic species, such as *P. cynomolgi* which share the same natural macaque hosts, have also been reported (6). Accurate detection of emerging zoonotic species such as *P. knowlesi* and *P. cynomolgi* in co-endemic areas with other human *Plasmodium* species infections is necessary to understand the geographical extent of zoonotic malaria transmission and to improve regional estimates of the disease burden (7). Improving national malaria control program detection and reporting of low-level *P. knowlesi* infections is also vital to demonstrate World Health Organization (WHO)-certified elimination for other non-zoonotic malaria species in Southeast Asia (8).

Conventional malaria diagnostic methods such as microscopy lack sensitivity and specificity for active surveillance of *P. knowlesi* due to common low-level sub-microscopic infections and an inability to accurately distinguish other morphologically similar *Plasmodium* species (7,9); notably *P. malariae* and the early ring stages of *P. falciparum* (10,11). Similarly, *P. cynomolgi* microscopically resembles *P. vivax* in human infections (12,13). Current malaria rapid diagnostic tests which detect circulating *Plasmodium* species antigens also remain insufficiently sensitive for *P. knowlesi* passive case detection at the low parasite counts able to produce symptomatic infections (14–16). Multiple molecular methods to detect *P. knowlesi* have been developed, including both quantitative and conventional qualitative PCR assays (7,17). However, systematic comparisons of the lower limit of detection (LOD) of these assays and exhaustive testing of *Plasmodium* species-specificity are currently lacking (7). The degree to which the LOD is enhanced with a prior reverse transcription (RT) step is not well characterised despite potential benefits in improving the detection of very low-level parasitemia symptomatic or asymptomatic zoonotic *Plasmodium* species infections (18). Specificity for *P. knowlesi* detection also ideally needs to be validated against other macaque zoonotic *Plasmodium* species, including *P. cynomolgi* (7).

To support an improved molecular surveillance workflow for detection of low-level zoonotic *Plasmodium* species infections (7,19), we evaluated an established *Plasmodium* genus and species-specific PCR assays with the inclusion of a reverse transcription step to enhance the LOD in both total nucleic acid preserved media and dried blood spot (DBS) collected samples.

## METHODS

### Clinical specimen collection and storage

Clinical venous whole blood samples and dried blood spots were collected prior to antimalarial treatment from individuals diagnosed with malaria by routine hospital microscopy, as part of an ongoing prospective malaria study in Sabah, Malaysia between April 2013 and May 2023. Adult healthy individuals were recruited as malaria-negative controls. Additional *P. vivax* clinical cases were enrolled from a prospective malaria study in western Indonesia between Jan 2022 and Aug 2023. Whole blood samples were collected in ethylenediaminetetraacetic acid (EDTA) and a small subset in DNA/RNA Shield^TM^ (Zymo Research, Irvine, CA, USA) before being frozen at -80°C at the time of enrolment. DBS were concurrently made using 20µL whole blood spotted on Whatman 3M filter paper and then stored in sealed bags with desiccant. A single donated *P. cynomolgi*-infected sample (20) obtained from a macaque host was frozen in glycerolyte and stored in liquid nitrogen prior to thawing, counting and immediately placing in DNA/RNA Shield^TM^.

### Microscopic parasite count quantification

Microscopic diagnosis of *Plasmodium* species was undertaken by experienced research microscopists using thick and thin Giemsa-stained blood films. Microscopic quantification of parasitemia (parasites per microlitre) was performed using thick blood smears calculated from the number of parasites per 200 white blood cells, multiplied by the individual patient’s total white cell count obtained from routine hospital laboratory flow cytometry (21).

### Total nucleic acid extraction and PCR amplification

Total nucleic acids were directly extracted from 200 μL of whole blood using a QIAamp® DNA Blood Mini Kit (QIAGEN, Cat. No. 51106), with DNA/RNA Shield^TM^ samples eluted in 50ul AE buffer to account for the preservative dilution factor. DNA and RNA extraction from DBS were carried out using an in-house method, established by Zainabadi *et al*, 2017

(22). Briefly, DBS equivalent to 40µl whole blood was incubated with 900µl lysis buffer at 65°C, with shaking at 250rpm for 90 minutes. Lysate was transferred to QIAamp® DNA Blood Mini Kit (QIAGEN, Cat. No. 51106) columns, washed with modified buffers, dried at 65°C for 10 minutes and eluted in 40µl buffer AE. The primers, annealing temperatures and/or probe sequences for each PCR assay are described in **Table 1**. Real-time PCR amplifications were performed on a QuantStudio^™^ 6 Flex (Applied Biosystems). Conventional PCR was performed on a DNA thermal cycler (Bio-Rad T100^™^ thermal cycler). The amplified nested PCR products were separated by electrophoresis using 2% agarose gels, stained by SYBR Safe™ (Invitrogen), and visualised on a UV transilluminator (Gel Doc XR+ imaging system, Bio-Rad). Each PCR amplification included a *Plasmodium* species positive and negative control and molecular weight standards (Applied Biosystems^TM^).

**Table 1.**
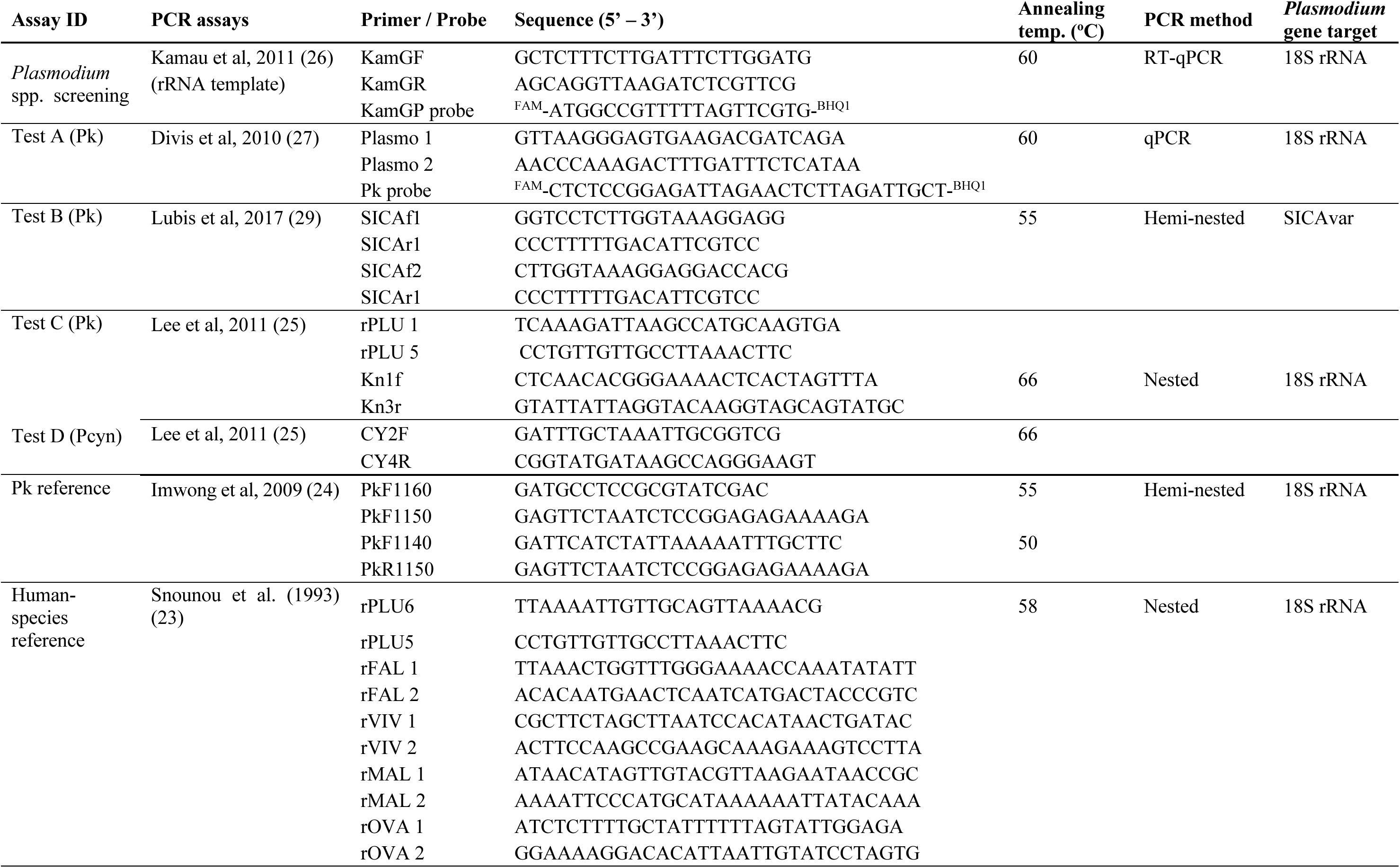
Primer sequences and annealing temperatures of *Plasmodium* genus and species-specific PCR asays.

### *Plasmodium* species confirmation using reference PCR

A validated reference PCR targeting the *Plasmodium* 18S rRNA genes was used to confirm the *Plasmodium* species using EDTA-whole blood clinical samples, consisting of an initial *Plasmodium* genus (hereafter abbreviated to *P*. genus) nest 1, followed by species-specific nest 2 for *P. falciparum*, *P. vivax*, *P. malariae*, and *P. ovale* spp. (23). The *P. knowlesi*-(24) and *P. cynomolgi*-specific (25) reference assays used also target 18S rRNA genes. The *P. knowlesi* reference assay has a previously reported high sensitivity (LOD less than 10 parasite genomes) when validated without reverse transcription (RT) against four *P. knowlesi* strains (Malayan, H, Philippine, and Hackeri); specificity was assessed against the major human *Plasmodium* species in addition to other simian *Plasmodium* species present in Southeast Asia including *P. cynomolgi*, *P. inui*, and *P. simiovale* (24).

### Selection of PCR assays for LOD evaluation

A reverse transcriptase real-time hydrolysis probe (RT-qPCR) assay designed to detect the *P. lasmodium* genus was evaluated for its utility in enhancing LOD (**Figure 1A**) (26). Three published *P. knowlesi* species-specific assays, which have not previously had their LODs evaluated using reverse transcription, were compared to the reference assay (**Figure 1B**). Test A is a qPCR assay (27) in current use in the State Public Health Laboratory (Makmal Kesihatan Awam; MKA) Sabah, Malaysia, for routine *P. knowlesi* malaria detection (28). Test B is a hemi-nested PCR targeting the multicopy *SICAvar* genes, previously validated for *P. knowlesi* detection against *P. falciparum*, *P. vivax*, *P. malariae*, and *P. ovale* spp., in addition to clinical *P. knowlesi* isolates from Sarawak, Malaysia (29). Test C is a nested conventional PCR assay targeting 18S rRNA genes specific for *P. knowlesi* that has been robustly validated against *P. vivax* and other zoonotic *Plasmodium* species (25).

**Figure 1.**
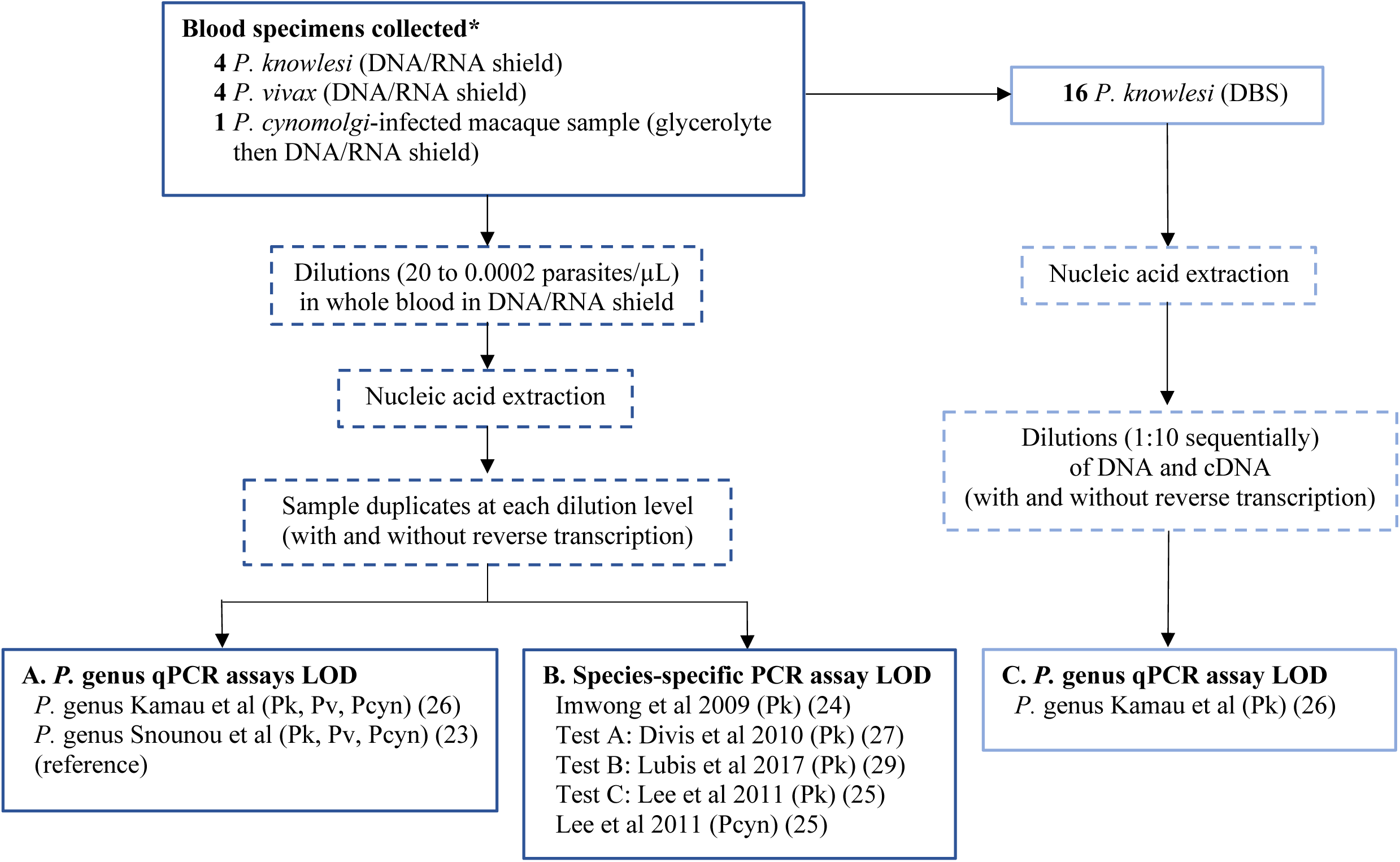
Limit of detection workflow for (A) *P.* genus, (B) *Plasmodium* species-specific, and (C) *P.* genus dried blood spot PCR assays. Abbreviations: DBS = dried blood spot; LOD = limit of detection; qPCR = real-time quantitative PCR; Pk *= P. knowlesi;* Pf *= P. falciparum,* Pv *= P. vivax;* Pcyn *= P. cynomolgi;* RT = reverse transcription

### Limit of detection evaluation for PCR assays with and without RT

The initial microscopy-quantified *Plasmodium* species infected samples collected in DNA/RNA Shield^TM^ were diluted with malaria-negative whole blood (also at the same manufacturer recommended DNA/RNA Shield^TM^ ratio) to prepare individual parasite count dilutions ranging between 20 to 0.0002 parasites/µL. Total nucleic acids from samples at each serial parasite count dilution were then extracted and duplicate aliquots prepared. High-capacity cDNA reverse transcription (Applied Biosystems^TM^, Thermo Fisher Scientific, MA, USA) was then performed on one aliquot from each pair. PCR was performed on the paired aliquots to detect *P.* genus by the qPCR assay (26), followed by species-specific assays for: *P. knowlesi* by the reference hemi-nested assay (24) and test assays A, B, C (25,27,29); *P. vivax* (23) and *P. cynomolgi* (25) by reference assays, in addition to the newly designed *P. vivax* primers. The LOD was expressed as the lowest parasite count per microlitre of whole blood detected by an individual PCR assay in both amplification replicates (**Figure 1C**).

### Limit of detection of *P.* genus qPCR assay for dried blood spots (DBS) with and without RT

The LOD for the *P.* genus qPCR assay was also evaluated using archived DBS from *P. knowlesi* clinical infections with and without RT (**Figure 1C**). DBS were stored individually after collection with dessicant at room temperature unexposed to light before processing. Nucleic acids were extracted from the DBS samples, with 10µl immediately reverse transcribed into cDNA. *P.* genus qPCR was conducted on serial 1:10 DNA and cDNA dilutions with the LOD calculated using the initial enumerated parasitemia divided by the corresponding dilution level.

### Evaluation of PCR assay specificity on clinical malaria samples

For the analysis of *P. knowlesi*, *P. falciparum*, and *P. vivax* clinical isolates, in addition to a *P. cynomolgi* macaque-derived isolate, all samples were individually tested using the *P.* genus qPCR assay and the three *P. knowlesi*-specific PCR assays. Results were compared against the reference PCR. Specificity for the *P.* genus assay was evaluated for malaria detection (any *Plasmodium* species) versus malaria-negative controls, and for species-specific assays using the corresponding *Plasmodium* species infection versus other *Plasmodium* species and malaria-negative controls combined. Clinical blood samples collected in EDTA likely had RNA degradation upon thawing; therefore, reverse transcription was not performed for this part of the analysis.

### Statistical analyses

Parasite counts for each clinical *Plasmodium* species infection were summarised using median and interquartile range (IQR). The median whole blood LOD was calculated with and without reverse transcription for the *P. knowlesi* and *P. vivax* isolates. To calculate the LOD fold-change, the LOD without RT was divided by the LOD with RT. One-way ANOVA was used to test for differences in parasite count distribution across *Plasmodium* species, followed by Student’s t-test for pairwise comparisons of log-transformed data. Results of PCR assays evaluated against reference PCR were defined as true positive (TP), false negative (FN), true negative (TN), and false positive (FP), enabling calculation of diagnostic sensitivity (TP/TP+FN) and specificity (TN/TN+FP) with exact binomial 95% confidence intervals. All statistical analyses were performed using Stata version 17.0 (StataCorp, Texas, USA).

## RESULTS

### Limit of detection of *P.* genus PCR assays

The LOD was performed on *P. knowlesi* (n=4), *P. vivax* (n=4) and *P. cynomolgi* (n=1) whole blood isolates. For the *P.* genus Kamau et al. qPCR assay, without reverse transcription, the median LOD to detect each individual *Plasmodium* species was 2 parasites/µL (**Table 2** and **Figure 2A**). With reverse transcription, the assay sensitivity for *P. knowlesi* improved with a LOD of ≤0.0002 parasites/µL for all four isolates (10,000-fold change). The LOD for both *P. vivax* and *P. cynomolgi* improved to 0.002 parasites/µL with RT (1,000-fold change); **Figure 2B**. In comparison, the reference Snounu *P.* genus assay had a slightly higher LOD for *P. vivax* and *P. cynomolgi* without RT (0.2 parasites/µL for both); however, with RT a less pronounced improvement in LOD was demonstrated at 0.02 and 0.01 parasites/µL, respectively.

**Figure 2.**
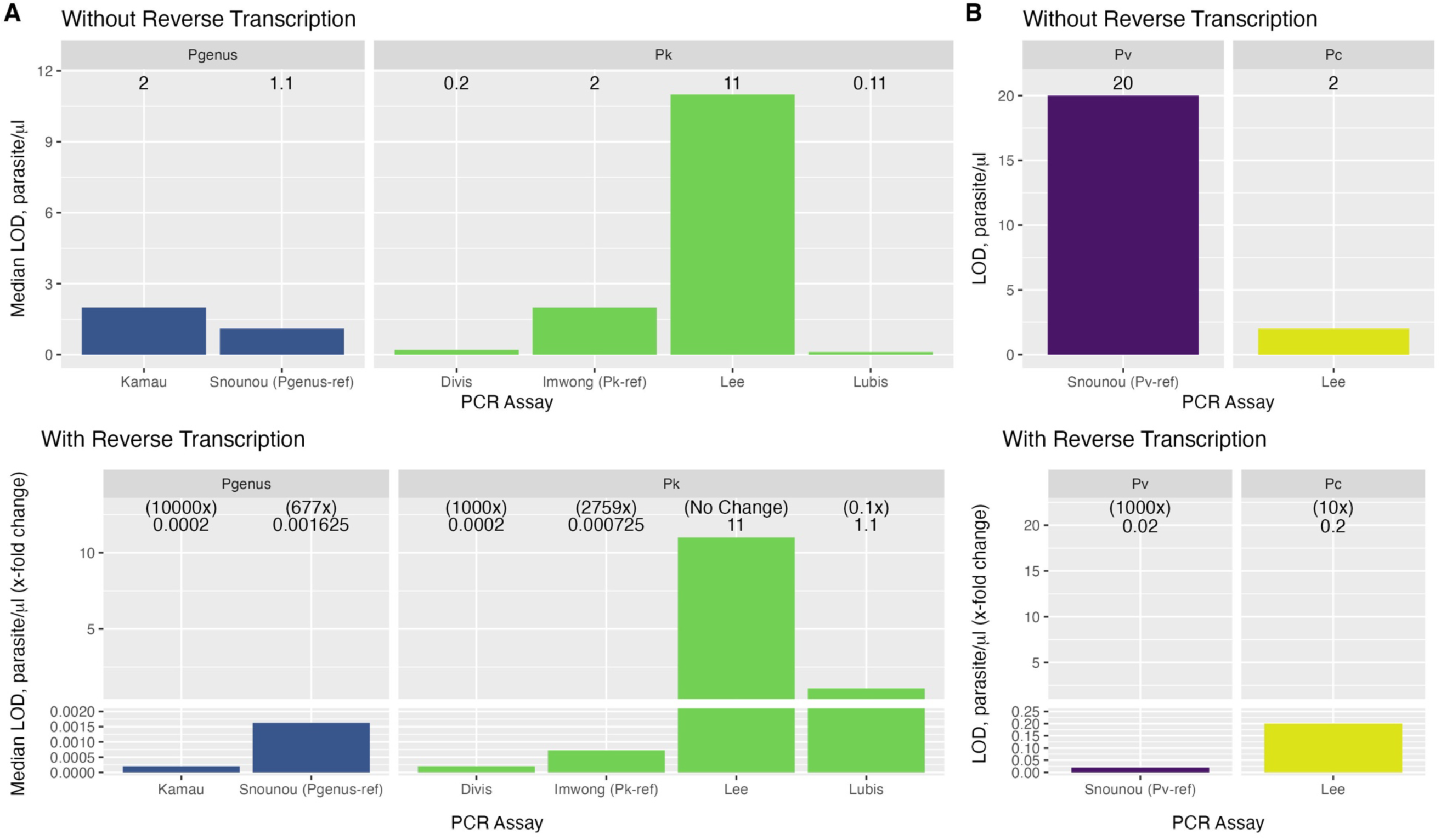
The median LOD and fold-change with and without reverse transcription for PCR assays to detect *Plasmodium* species. **(A)** The median LOD (parasites/µL) for *P.* genus assays using 6 clinical isolates (*P. knowlesi*=4, *P. vivax*=1, *P. cynomolgi*=1) and *P. knowlesi*-specific assays using 4 clinical isolates; **(B)** The LOD (parasites/µL) for *P. vivax* (n=1) and *P. cynomolgi* (n=1) specific assays. Abbreviations: Pgenus, *Plasmodium* genus; Pk, *P. knowlesi*; Pv, *P. vivax;* Pc, *P. cynomolgi*

**Table 2.**
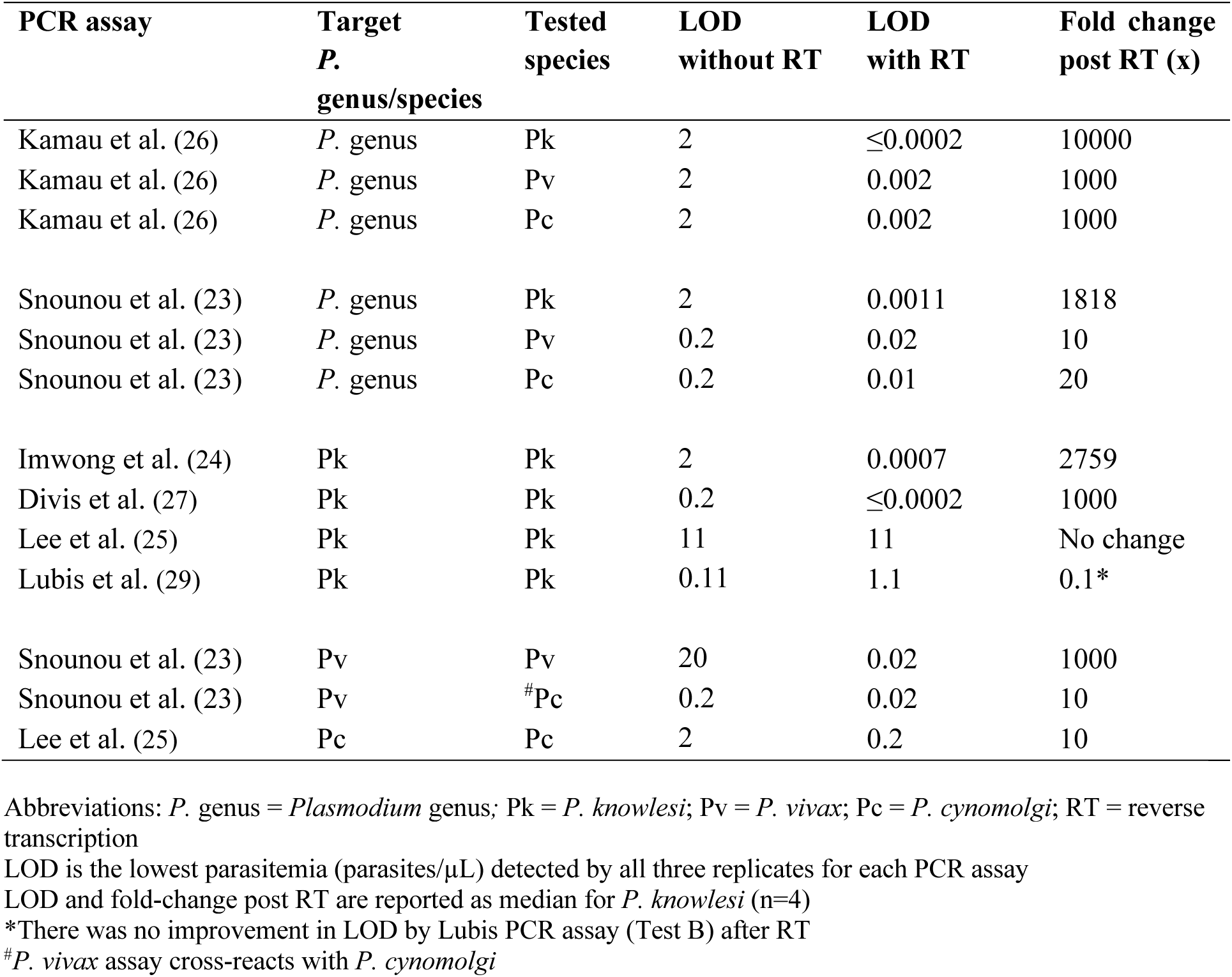
Limit of detection (LOD) of *Plasmodium* genus and species-specific PCR assays.

### Limit of detection of *Plasmodium* species*-*specific PCR assays

For the *P. knowlesi*-specific assays, the median LOD without and with reverse transcription was 2 and 0.0007 parasites/µL, respectively, for the Imwong et al. reference assay (2759-fold change); 0.2 and 0.0002 parasites/µL, respectively, for Test A (1000-fold); 0.11 and 1.1 parasites/µL, respectively, for Test B (no improvement); and 11 parasites/µL for both (no change) for Test C (**Table 2 and Figure 2A)**.

Without reverse transcription, the LODs using species-specific reference assays were 2 parasites/µL for *P. cynomolgi* and 20 parasites/µL for *P. vivax*. With reverse transcription, the LODs were 0.2 (10-fold change) and 0.02 (1000-fold change) for *P. cynomolgi* and *P. vivax* respectively. However, additional testing of the *P. cynomolgi* isolate using the reference *P. vivax*-targeted rVIV1/rVIV2 primers (23) amplified this target from the macaque-origin *P. cynomolgi* infection both without (0.2 parasites/µL) and with reverse transcription (0.02 parasites/µL), producing a false-positive *P. vivax* result (**Table 2 and Figure 2B**).

The LOD with reverse transcription of the *P.* genus assay was comparable to the LOD of the best performing species-specific assay for *P. knowlesi* detection (≤0.0002 parasites/µL for both). In contrast, the *P.* genus assay had a superior LOD compared to the reference species-specific assays for *P. vivax* (0.002 vs 0.02 parasites/µL respectively) and *P. cynomolgi* (0.002 vs 0.2 parasites/µL respectively).

### Limit of detection of *P.* genus qPCR for dried blood spots (DBS)

The median LOD between DNA and cDNA generated from dried blood spots (DBS) for 4 *P. knowlesi* extracted samples collected 8 months prior to evaluation of the *P.* genus Kamau 2011 assay without and with RT was 19.86 and 0.08 parasites/uL, respectively (249-fold change); **Table 3**. Archived DBS samples (n=12) collected more than 6 years (up to 11 years) prior to extraction demonstrated a similar LOD with and without RT (median 20 parasites/µL).

**Table 3.**
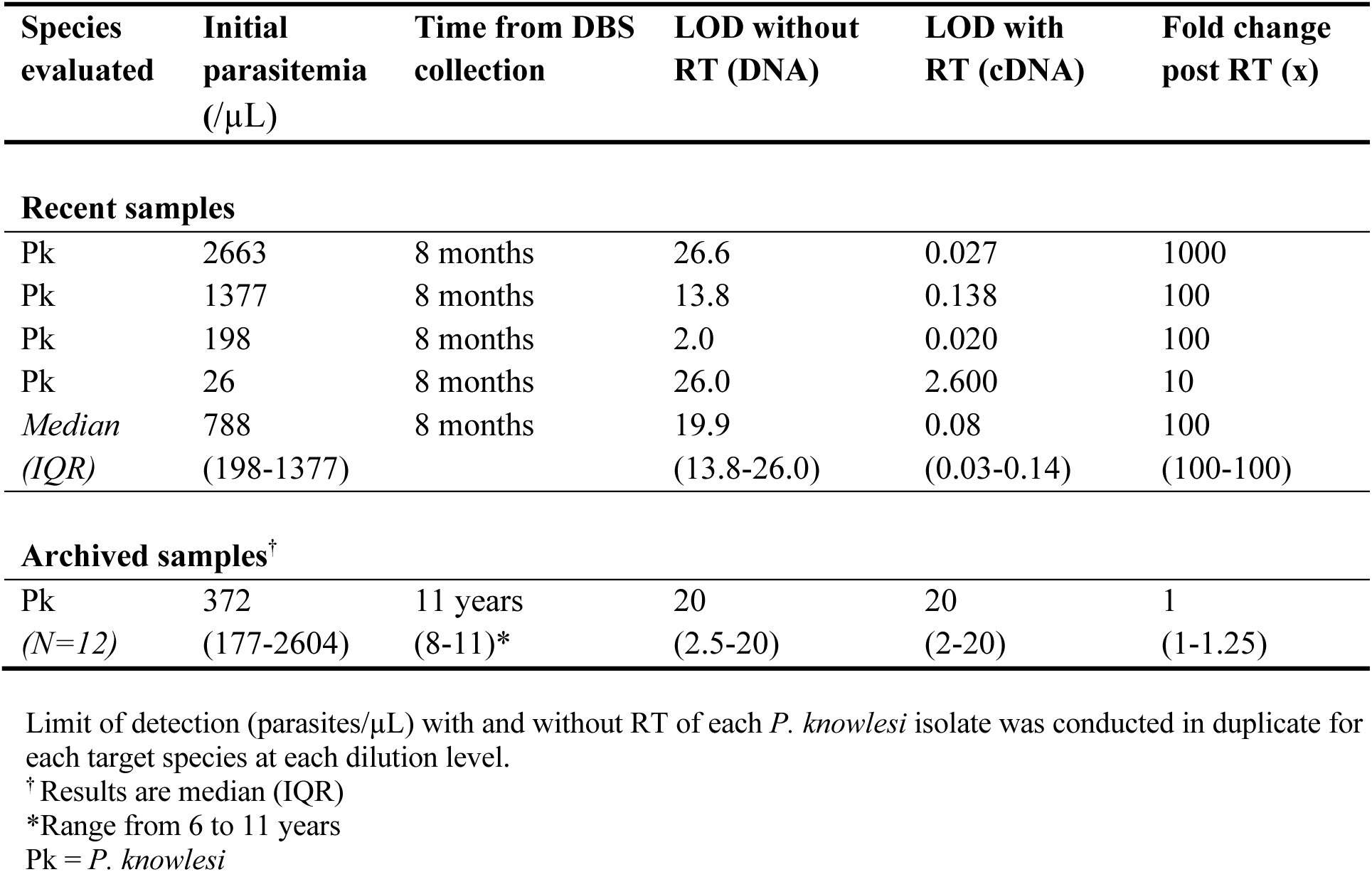
Limit of detection of the 18S rRNA *P.* genus qPCR assay for *P. knowlesi* dried blood spot samples.

### Specificity of *P.* genus and individual *Plasmodium* species PCR assays

A total of 239 samples were included in the clinical evaluation of test specificity without RT, consisting of 96 *P. knowlesi*, 50 *P. vivax,* 44 *P. falciparum*, and 1 *P. cynomolgi* infected samples, and 48 malaria-negative controls. The median parasitemia was 1,957/µL (IQR 261-5,762; range 27-210,100 parasites/µL) for *P. knowlesi*, 3,246/µL (IQR 1,588-7,306; range 77-20,064 parasites/µL) for *P. vivax*, and 14,015/µL (IQR 2,193-33,413; range 34-297,000 parasites/µL) for *P. falciparum*.

The *P.* genus qPCR screening assay was both 100% specific and sensitive for the detection of *Plasmodium* species overall compared to the reference PCR, with all 48 samples from uninfected controls confirmed as malaria-negative.

*P. knowlesi*-assays for Tests A (27) and B (29) correctly identified all 96 *P. knowlesi* and 95 non-*P. knowlesi* samples, resulting in 100% specificity and sensitivity. Test C (25) was negative for a single *P. knowlesi* isolate with a parasite count of 1,535 parasites/µL, resulting in 99% (95% CI 94.3-100.0) sensitivity and 100% specificity. The reference assays for *P. falciparum* and *P. vivax* (23), and for *P. cynomolgi* (25) were negative for all *P. knowlesi* clinical isolates tested (**Table 2**).

## DISCUSSION

Malaria-susceptible countries in most of Southeast Asia, including those approaching or achieving WHO elimination of major human-only *Plasmodium* species, remain at-risk for zoonotic malaria transmission (30,31). Understanding regional heterogeneity in *P. knowlesi* transmission intensity and disease morbidity will require the selective deployment of highly sensitive and specific molecular detection tools for both diagnostic and surveillance purposes (19). Our major finding demonstrated that the use of a reverse-transcription step after extraction of preserved total nucleic acids in clinical samples considerably improves the limit of detection of both the selected *P.* genus RT-qPCR screening assay, and *P. knowlesi*-specific assays (reference and Test A; both >1000-fold) by additionally amplifying ribosomal RNA sequences. The enhanced limit of detection was consistent across both field-stable DNA/RNA Shield^TM^ samples and to a lesser degree in recent (although not older) dried blood spots. The second key finding of this study was the excellent performance of the *P.* genus screening assay, originally developed and validated for use in an African context for *P. falciparum* (26), to detect previously unvalidated species including *P. knowlesi*, *P. cynomolgi* and *P. vivax*. The specificity of each of the *P. knowlesi*-targeted assays to exclude *P. cynomolgi* and non-zoonotic *Plasmodium* species was confirmed to be excellent. Together these findings highlight the potential utility of incorporating these assays in a molecular surveillance approach to detect both human and zoonotic *Plasmodium* species that are well below the reported parasite count detection limits for current conventional PCR, loop-mediated isothermal amplification, microscopy or parasite lactate-dehydrogenase-based rapid diagnostic tests (19).

The community-based detection of submicroscopic *P. knowlesi* and *P. cynomolgi* infections, both symptomatic or asymptomatic, requires ultrasensitive molecular tools to understand the true extent of population-level transmission (19). Recent studies in areas of both Peninsular Malaysia and the East Malaysian state of Sarawak have reported human infections in local communities living in or near forested areas with other macaque malaria species, including *P. inui*, *P. coatneyi, P. fieldi* and possibly *P. simiovale,* in addition to *P. knowlesi* and *P. cynomolgi* (32). It is unclear whether or to what extent these low-level zoonotic infections may facilitate onward transmission to humans (33,34), as occurs with low parasitemia *P. falciparum* and *P. vivax* infections (35,36), although sustained human-to-human transmission of *P. knowlesi* has not been evident to date(25,37).

The selection of the major RT-qPCR *P.* genus screening assay aimed to maximise the detection limits for low-level zoonotic *Plasmodium* species infections due to the known high multicopy number (5 to 10 copies per genome depending on the *Plasmodium* species) of the 18S rRNA target (38), in addition to amplification of both the A- and S-type genes and their RNA transcripts (26). The excellent LOD demonstrated with the *P.* genus RT-qPCR assay in the current study of <0.0002 parasites/µL for *P. knowlesi* detection is consistent with a previously reported extremely low LOD of ∼0.0004 parasites/µL for clinical *P. falciparum* samples (26). DNA-concentrated packed red blood cell samples have been demonstrated to further improve sensitivity for *P. falciparum* detection in population-based malaria prevalence surveys in elimination areas of Thailand (39). Comparable performance to this *P.* genus RT-qPCR assay was reported with a separately established ultrasensitive quantitative PCR method (uPCR) with a limit of detection of 0.022 parasites/µL (39), however, a major advantage of the reverse transcriptase qPCR assay is the requirement for comparatively lower blood volumes (18). Interestingly, in the current study the conventional PCR *P.* genus reference assay also demonstrated a large increase in analytical sensitivity after reverse transcription (∼1800-fold) and may provide a more cost-effective option compared to qPCR for surveillance purposes. DNA/RNA Shield^TM^ was selected as the preferred blood collection preservation method over other media due to its reported ability to stabilise DNA/RNA at ambient temperatures in field settings and compatibility with most DNA and RNA purification kits for subsequent high-throughput workflows including reverse transcription (40).

To date, only a few studies have incorporated reverse transcription in the molecular detection of *Plasmodium* species (41,42). The reverse transcription step in the present study improved the analytical sensitivity of our selected assays to detect zoonotic *P. knowlesi*, *P. cynomolgi* and other human malaria infections by up to 10,000-fold (*Plasmodium* genus), 2759-fold (*P. knowlesi*), 1000-fold (*P. vivax*) and 10-fold (*P. cynomolgi*), respectively. The *P. knowlesi*-specific hemi-nested reference assay with reverse transcription demonstrated a comparable limit of detection to the qPCR Test A (which requires an expensive real-time hydrolysis probe), and was superior to both Test B targeting *SICavar* and Test C targeting the 18S rRNA gene. Without reverse transcription, the lowest limit of detection for *P. knowlesi* was seen with Test B, suggesting that the multiple chromosomal copies of the variant antigen *SICAvar* provide equivalent or better signal amplification than the detection of transcripts from whichever of these gene copies is activated in any particular parasite cell in the peripheral blood. Constraints on the widespread use of reverse transcription include the additional cost, laboratory time, and the usual rapid degradation of RNA molecules in field or laboratory settings. However, the degree of RNA amplification with reverse transcription was aided in our study by collecting blood samples in room-temperature stable RNA preservation media suitable for field-based surveillance, which also allows other potential downstream pathophysiological or transcriptomic analyses dependent on pathogen or host RNA transcripts.

The low reported limit of detection for the RT-qPCR *P.* genus assay conducted on DBS in this study (∼0.08 parasites/µL with reverse transcription) suggests this type of sample collection would also enable the identification of a large proportion of submicroscopic and/or asymptomatic infections. DBS collection is logistically a more feasible, inexpensive and acceptable option particularly for asymptomatic or younger participants (given the need for fingerprick blood collection rather than venepuncture) for large-scale malaria surveillance surveys. However, the reverse transcription step only improved the LOD in DBS samples that were collected within 8 months; older DBS stored in recommended conditions for more than 6 years did not provide any improvement in the LOD with and without RT due to likely degradation of RNA. Regardless, the *P.* genus LOD of around 2 to 20 parasites/µL without RT for DBS samples remains encouraging as a first-line option for surveillance screening purposes, although the use of DBS would require further evaluation with *Plasmodium* species-specific PCR assay differentiation.

The current study confirmed previous findings detailing cross-reactivity between the nested PCR primers for *P. vivax* (rVIV1/rVIV2) with *P. cynomolgi* (13). A single mismatch in the 30 nucleotides of the rVIV2 primer sequences was reported to cross-amplify *P. cynomolgi* isolates (13). The nested *P. vivax-*specific assay using the same primers rVIV1/rVIV2 designed to target the 18S rRNA gene also amplified *P. cynomolgi* in our LOD analysis (23). The separate *P. cynomolgi* primers remained highly specific and did not erroneously amplify *P. vivax* or other closely related *Plasmodium* species DNA. In practice, the concurrent use of both assays would enable accurate identification of a *P. vivax* mono-infection, however, this approach would not be able to differentiate a *P. vivax/P. cynomolgi* co-infection from a *P. cynomolgi* mono-infection. Mis-identification of symptomatic *P. cynomolgi* infections as *P. vivax* would not result in inappropriate treatment, given both have a latent hypnozoite liver life-stage requiring additional radical cure with primaquine. Most other commonly used single-round multiplex (43) and qPCR assays (44) containing *P. vivax-*specific targets have also not been validated against isolates of *P. cynomolgi* or other closely related macaque *Plasmodium* species. However, a variety of sequencing approaches of targeted gene amplicons including mitochondrial COX1 and cytochrome b, *SICAvar* and SSU 18S rRNA followed by sequencing and reference alignment have been used to confirm unknown or mixed zoonotic *Plasmodium* species infections following initial ambiguous PCR results (45,46).

A limitation of this study was the inability to validate submicroscopic clinical *P. knowlesi* infections and other zoonotic species such as *P. fieldi*, *P. inui* and *P. coatneyi* for which samples were not available. Due to the increasing number of published *P. knowlesi* assays, we were not able to evaluate other *P. knowlesi* assays of possibly comparable performance within our selected workflow. We were also unable to evaluate mixed infections of *P. knowlesi*, *P. vivax* and *P. falciparum* despite these cases being reported in certain areas such as Indonesia (29) and Vietnam (47). Due to sample availability, we were also restricted to only a single *P. cynomolgi* sample to determine the LOD of the *P. cynomolgi-*specific assay (25). The discrepancy between the LOD for the *P.* genus qPCR screening assay and the species-specific assay for *P. cynomolgi* detection may mean a proportion of very low-level *P. cynomolgi* infections are unable to be identified beyond a *P.* genus threshold using the current protocol.

## CONCLUSIONS

The *Plasmodium* genus reverse transcriptase qPCR assay can provide highly sensitive screening for zoonotic and human malaria, including for submicroscopic infections in at-risk populations in endemic areas. The use of this molecular surveillance protocol for either whole blood or DBS collected samples in understudied areas of Southeast Asia would enable improved understanding of the regional disease burden and transmission dynamics of zoonotic malaria. Enhanced molecular tools and future iterative improvements to conventional surveillance protocols are especially critical as Southeast Asia continues to exert considerable public health efforts towards human malaria elimination despite the challenge of additional zoonotic *Plasmodium* species infections at an expanding human-animal-interface.

## Data Availability

All data produced in the present work are contained in the manuscript

## DECLARATIONS

### Ethics approval and consent to participate

Sample collection and diagnostic evaluation were approved as part of prospective malaria studies by the Medical Research and Ethics Committee (MREC), Ministry of Health Malaysia (NMRR-10-754-6684, NMRR-19-4109-52172 and NMRR-19-3229-49967), Universitas Sumatera Utara, Indonesia (#723/KEP/USU/2021) and by the Menzies School of Health Research, Australia (HREC-2010-1431 and HREC-2022-4417) in accordance with all applicable Federal and other regulations governing the protection of human subject research.

### Competing interests

The authors declare that they have no competing interests.

### Funding

This work was supported by the ZOOMAL project, funded through the Australian Centre for International Agricultural Research and Indo-Pacific Centre for Health Security, DFAT, Australian Government (#LS-2019-116), the National Institutes of Health, USA (#R01AI160457-01), and DOD-DHA-Global Emerging Infections Surveillance program project P0097_22_N2.). Funding support was also through the National Health and Medical Research Council, Australia (Grant Numbers #1037304 and #1045156, fellowship to NMA [#1042072], Emerging Leadership 2 Investigator Grants to MJG [#2017436] and BEB [#2016792]), and the Ministry of Health, Malaysia (#BP00500/117/1002) awarded to GSR.

### Disclaimers

The views expressed are those of the author(s) and do not necessarily reflect the official policy or position of the Department of the Navy, Department of Defense, nor the U.S. Government. Copyright Statement: NC (LCDR, MSC, USN) & AL (CAPT, MC, USN) are military service members and employees of the U.S. Government. This work was prepared as part of their official duties at U.S. Naval Medical Research Unit INDO PACIFIC. Title 17 U.S.C. §105 provides that ‘Copyright protection under this title is not available for any work of the United States Government.’ Title 17 U.S.C. §101 defines a U.S. Government work as a work prepared by a military service member or employee of the U.S. Government as part of that person’s official duties.

### Authors’ contributions

Conceptualization, funding acquisition, methodology and resources: MG, NA, KB, KP, INL, RN, CP, NC, AL. Sample processing and conduct of assays: KP and PK. Data analyses: KB, MG, KP. Initial manuscript preparation and writing: KB, MG. All authors read and approved the final manuscript.

## Acknowledgements

We thank the study participants, the IDSKKS malaria research team (Mohd Rizan Osman, Danshy Alaza, Azielia Elastiqah, Sitti Saimah binti Sakam), and Bruce Russell of the Department of Microbiology & Immunology, University of Otago, Dunedin, New Zealand, for providing the *P. cynomolgi* isolate. We thank the Director General of Health Malaysia for the permission to publish this article.

## Notes

### Competing Interest Statement

The authors have declared no competing interest.

## REFERENCES

1. Moyes CL, Henry AJ, Golding N, Huang Z, Singh B, Baird JK, et al. Defining the Geographical Range of the *Plasmodium knowlesi* Reservoir. PLoS Neglected Trop Dis. 2014;8(3):e2780.

2. Cuenca PR, Key S, Jumail A, Surendra H, Ferguson HM, Drakeley C, et al. Epidemiology of the zoonotic malaria *Plasmodium knowlesi* in changing landscapes. Adv Parasit. 2021;113:225–86.

3. Barber BE, William T, Grigg MJ, Menon J, Auburn S, Marfurt J, et al. A Prospective Comparative Study of Knowlesi, Falciparum, and Vivax Malaria in Sabah, Malaysia: High Proportion With Severe Disease From *Plasmodium knowlesi* and *Plasmodium vivax* But No Mortality With Early Referral and Artesunate Therapy. Clin Infect Dis. 2013 Feb;56(3):383–97.

4. Daneshvar C, Davis TME, Cox-Singh J, Rafa’ee MZ, Zakaria SK, Divis PCS, et al. Clinical and laboratory features of human *Plasmodium knowlesi* infection. Clinical Infectious Diseases [Internet]. 2009 Sep 15;49(6):852–60. Available from: http://cid.oxfordjournals.org/lookup/doi/10.1086/605439

5. Grigg MJ, William T, Barber BE, Rajahram GS, Menon J, Schimann E, et al. Age-Related Clinical Spectrum of *Plasmodium knowlesi* Malaria and Predictors of Severity. Clin Infect Dis. 2018 Aug 1;67(3):350–9.

6. Kotepui M, Masangkay FR, Kotepui KU, Milanez GDJ. Preliminary review on the prevalence, proportion, geographical distribution, and characteristics of naturally acquired *Plasmodium cynomolgi* infection in mosquitoes, macaques, and humans: a systematic review and meta-analysis. BMC Infect Dis. 2021;21(1):259.

7. Grigg MJ, Lubis IN, Tetteh KKA, Barber BE, William T, Rajahram GS, et al. *Plasmodium knowlesi* detection methods for human infections—Diagnosis and surveillance. In: Advances in Parasitology [Internet]. Academic Press; 2021. p. 77–130. (Drakeley C, editor. Advances in Parasitology; vol. 113). Available from: https://www.sciencedirect.com/science/article/pii/S0065308X21000270

8. WHO. Malaria Policy Advisory Group (MPAG) meeting (March 2022) [Internet]. 2022 Apr. Available from: https://www.who.int/publications/i/item/9789240048430

9. Barber BE, William T, Grigg MJ, Yeo TW, Anstey NM. Limitations of microscopy to differentiate *Plasmodium* species in a region co-endemic for *Plasmodium falciparum*, Plasmodium vivax and Plasmodium knowlesi. Malaria J. 2013 Jan 8;12(1):8.

10. Lee KS, Cox-Singh J, Singh B. Morphological features and differential counts of *Plasmodium knowlesi* parasites in naturally acquired human infections. Malaria J. 2009 Apr 23;8(1):73–73.

11. Coutrier FN, Tirta YK, Cotter C, Zarlinda I, González IJ, Schwartz A, et al. Laboratory challenges of *Plasmodium* species identification in Aceh Province, Indonesia, a malaria elimination setting with newly discovered P. knowlesi. PLoS Neglected Trop Dis [Internet]. 2018;12(11):e0006924. Available from: 10.1371/journal.pntd.0006924

12. Hartmeyer GN, Stensvold CR, Fabricius T, Marmolin ES, Hoegh SV, Nielsen HV, et al. *Plasmodium cynomolgi* as Cause of Malaria in Tourist to Southeast Asia, 2018. Emerging infectious diseases. 2019 Oct;25(10):1936–9.

13. Ta TH, Hisam S, Lanza M, Jiram AI, Ismail N, Rubio JM. First case of a naturally acquired human infection with *Plasmodium cynomolgi*. Malaria J. 2014;13(1):68.

14. Barber BE, William T, Grigg MJ, Piera K, Yeo TW, Anstey NM. Evaluation of the Sensitivity of a pLDH-Based and an Aldolase-Based Rapid Diagnostic Test for Diagnosis of Uncomplicated and Severe Malaria Caused by PCR-Confirmed *Plasmodium knowlesi*, *Plasmodium falciparum*, and *Plasmodium vivax*. J Clin Microbiol. 2013 Apr 1;51(4):1118– 23.

15. Grigg MJ, William T, Barber BE, Parameswaran U, Bird E, Piera K, et al. Combining Parasite Lactate Dehydrogenase-Based and Histidine-Rich Protein 2-Based Rapid Tests To Improve Specificity for Diagnosis of Malaria Due to *Plasmodium knowlesi* and Other *Plasmodium* Species in Sabah, Malaysia. J Clin Microbiol. 2014 Apr;52(6):2053–60.

16. Tan AF, Sakam SS binti, Rajahram GS, William T, Isnadi MFAR, Daim S, et al. Diagnostic accuracy and limit of detection of ten malaria parasite lactate dehydrogenase-based rapid tests for *Plasmodium knowlesi* and *P. falciparum*. Front Cell Infect Microbiol [Internet]. 2022;12:1023219. Available from: https://www.frontiersin.org/articles/10.3389/fcimb.2022.1023219

17. Lee PC, Chong ETJ, Anderios F, Lim YA, Chew CH, Chua KH. Molecular detection of human *Plasmodium* species in Sabah using PlasmoNex^TM^ multiplex PCR and hydrolysis probes real-time PCR. Malaria Journal. 2015 Jan 28;14(1):28.

18. Christensen P, Bozdech Z, Watthanaworawit W, Renia L, Malleret B, Ling C, et al. Reverse transcription PCR to detect low density malaria infections. Wellcome Open Res. 2021;6:39.

19. Anstey NM, Grigg MJ. Zoonotic Malaria: The Better You Look, the More You Find. J Infect Dis [Internet]. 2019;219(5):679–81. Available from: https://www.ncbi.nlm.nih.gov/pubmed/30295775

20. Christensen P, Racklyeft A, Ward KE, Matheson J, Suwanarusk R, Chua ACY, et al. Improving in vitro continuous cultivation of *Plasmodium cynomolgi*, a model for *P. vivax*. Parasitol Int. 2022;89:102589.

21. WHO. Microscopy for the detection, identification and quantification of malaria parasites on stained thick and thin blood films in research settings. World Health Organisation [Internet]. 2015; Available from: https://apps.who.int/iris/handle/10665/163782

22. Zainabadi K, Adams M, Han ZY, Lwin HW, Han KT, Ouattara A, et al. A novel method for extracting nucleic acids from dried blood spots for ultrasensitive detection of low-density *Plasmodium falciparum* and *Plasmodium vivax* infections. Malar J [Internet]. 2017;16(1):377. Available from: 10.1186/s12936-017-2025-3

23. Snounou G, Viriyakosol S, Jarra W, Thaithong S, Brown KN. Identification of the four human malaria parasite species in field samples by the polymerase chain reaction and detection of a high prevalence of mixed infections. Molecular and biochemical parasitology. 1993 Apr;58(2):283–92.

24. Imwong M, Tanomsing N, Pukrittayakamee S, Day NPJ, White NJ, Snounou G. Spurious Amplification of a *Plasmodium vivax* Small-Subunit RNA Gene by Use of Primers Currently Used To Detect *P. knowlesi*. J Clin Microbiol [Internet]. 2009;47(12):4173–5. Available from: https://www.ncbi.nlm.nih.gov/pubmed/19812279

25. Lee KS, Divis PCS, Zakaria SK, Matusop A, Julin RA, Conway DJ, et al. *Plasmodium knowlesi*: Reservoir Hosts and Tracking the Emergence in Humans and Macaques. Kazura JW, editor. PLoS Pathog. 2011;7(4):e1002015.

26. Kamau E, Tolbert LS, Kortepeter L, Pratt M, Nyakoe N, Muringo L, et al. Development of a Highly Sensitive Genus-Specific Quantitative Reverse Transcriptase Real-Time PCR Assay for Detection and Quantitation of *Plasmodium* by Amplifying RNA and DNA of the 18S rRNA Genes. J Clin Microbiol [Internet]. 2011;49(8):2946–53. Available from: https://www.ncbi.nlm.nih.gov/pubmed/21653767

27. Divis PC, Shokoples SE, Singh B, Yanow SK. A TaqMan real-time PCR assay for the detection and quantitation of *Plasmodium knowlesi*. Malar J [Internet]. 2010;9(1):344. Available from: https://www.ncbi.nlm.nih.gov/pubmed/21114872

28. Nuin NA, Tan AF, Lew YL, Piera KA, William T, Rajahram GS, et al. Comparative evaluation of two commercial real-time PCR kits (QuantiFast^TM^ and abTES^TM^) for the detection of *Plasmodium knowlesi* and other *Plasmodium* species in Sabah, Malaysia. Malaria J. 2020 Aug;19(1):306.

29. Lubis IND, Wijaya H, Lubis M, Lubis CP, Divis PCS, Beshir KB, et al. Contribution of *Plasmodium knowlesi* to Multispecies Human Malaria Infections in North Sumatera, Indonesia. J Infect Dis [Internet]. 2017;215(7):1148–55. Available from: https://www.ncbi.nlm.nih.gov/pubmed/28201638

30. Shearer FM, Huang Z, Weiss DJ, Wiebe A, Gibson HS, Battle KE, et al. Estimating Geographical Variation in the Risk of Zoonotic *Plasmodium knowlesi* Infection in Countries Eliminating Malaria. Churcher TS, editor. PLoS Neglected Trop Dis [Internet]. 2016;10(8):e0004915. Available from: 10.1371/journal.pntd.0004915

31. Tobin RJ, Harrison LE, Tully MK, Lubis IND, Noviyanti R, Anstey NM, et al. Updating estimates of *Plasmodium knowlesi* malaria risk in response to changing land use patterns across Southeast Asia. PLOS Neglected Trop Dis. 2024;18(1):e0011570.

32. Yap NJ, Hossain H, Nada-Raja T, Ngui R, Muslim A, Hoh BP, et al. Natural Human Infections with *Plasmodium cynomolgi*, *P. inui*, and 4 other Simian Malaria Parasites, Malaysia - Volume 27, Number 8—August 2021 - Emerging Infectious Diseases journal - CDC. Emerg Infect Dis [Internet]. 2021;27(8):2187–91. Available from: https://www.ncbi.nlm.nih.gov/pubmed/34287122

33. Fornace KM, Nuin NA, Betson M, Grigg MJ, William T, Anstey NM, et al. Asymptomatic and Submicroscopic Carriage of *Plasmodium knowlesi* Malaria in Household and Community Members of Clinical Cases in Sabah, Malaysia. J Infect Dis. 2016 Mar 1;213(5):784–7.

34. Noordin NR, Lee PY, Bukhari FDM, Fong MY, Hamid MHA, Jelip J, et al. Prevalence of Asymptomatic and/or Low-Density Malaria Infection among High-Risk Groups in Peninsular Malaysia. Am J Trop Med Hyg. 2020;103(3):1107–10.

35. Schneider P, Bousema JT, Gouagna LC, Otieno S, Vegte-Bolmer MV de, Omar SA, et al. Submicroscopic *Plasmodium falciparum* gametocyte densities frequently result in mosquito infection. American Journal of Tropical Medicine and Hygiene. 2007;76(3):470–4.

36. Almeida ACG, Kuehn A, Castro AJM, Vitor-Silva S, Figueiredo EFG, Brasil LW, et al. High proportions of asymptomatic and submicroscopic *Plasmodium vivax* infections in a peri-urban area of low transmission in the Brazilian Amazon. Parasites Vectors. 2018;11(1):194.

37. Fornace KM, Topazian HM, Routledge I, Asyraf S, Jelip J, Lindblade KA, et al. No evidence of sustained nonzoonotic *Plasmodium knowlesi* transmission in Malaysia from modelling malaria case data. Nat Commun. 2023;14(1):2945.

38. Mercereau-Puijalon O, Barale JC, Bischoff E. Three multigene families in *Plasmodium* parasites: facts and questions. Int J Parasitol [Internet]. 2002;32(11):1323–44. Available from: https://www.sciencedirect.com/science/article/pii/S002075190200111X

39. Imwong M, Hanchana S, Malleret B, Renia L, Day NPJ, Dondorp A, et al. High-throughput ultrasensitive molecular techniques for quantifying low-density malaria parasitemias. Gilligan PH, editor. Journal of clinical microbiology [Internet]. 2014 Sep;52(9):3303–9. Available from: https://www.ncbi.nlm.nih.gov/pmc/articles/PMC4313154/pdf/zjm3303.pdf

40. Eberhardt E, Hendrickx R, Kerkhof MV den, Monnerat S, Alves F, Hendrickx S, et al. Comparative evaluation of nucleic acid stabilizing reagents for RNA- and DNA-based Leishmania detection in blood as proxy for visceral burdens. J Microbiol Methods. 2020;173:105935.

41. Adams M, Joshi SN, Mbambo G, Mu AZ, Roemmich SM, Shrestha B, et al. An ultrasensitive reverse transcription polymerase chain reaction assay to detect asymptomatic low-density *Plasmodium falciparum* and *Plasmodium vivax* infections in small volume blood samples. Malar J. 2015 Dec 23;14(1):520.

42. Bousema T, Banman SL, Taylor BJ, Rijpma SR, Morlais I, Yanow SK, et al. A Direct from Blood Reverse Transcriptase Polymerase Chain Reaction Assay for Monitoring Falciparum Malaria Parasite Transmission in Elimination Settings. Am J Trop Med Hyg. 2017;97(2):533–43.

43. Padley D, Moody AH, Chiodini PL, Saldanha J. Use of a rapid, single-round, multiplex PCR to detect malarial parasites and identify the species present. Ann Trop Med Parasitol [Internet]. 2003;97(2):131–7. Available from: 10.1179/000349803125002977

44. Rougemont M, Saanen MV, Sahli R, Hinrikson HP, Bille J, Jaton K. Detection of Four *Plasmodium* Species in Blood from Humans by 18S rRNA Gene Subunit-Based and Species-Specific Real-Time PCR Assays. J Clin Microbiol [Internet]. 2004;42(12):5636–43. Available from: https://journals.asm.org/doi/abs/10.1128/JCM.42.12.5636-5643.2004

45. Imwong M, Madmanee W, Suwannasin K, Kunasol C, Peto TJ, Tripura R, et al. Asymptomatic Natural Human Infections With the Simian Malaria Parasites *Plasmodium cynomolgi* and *Plasmodium knowlesi*. J Infect Dis [Internet]. 2019;219(5):695–702. Available from: https://www.ncbi.nlm.nih.gov/pubmed/30295822

46. Putaporntip C, Kuamsab N, Seethamchai S, Pattanawong U, Rojrung R, Yanmanee S, et al. Cryptic *Plasmodium inui* and *Plasmodium fieldi* Infections Among Symptomatic Malaria Patients in Thailand. Clin Infect Dis. 2021;75(5):805–12.

47. Marchand RP, Culleton R, Maeno Y, Quang NT, Nakazawa S. Co-infections of *Plasmodium knowlesi, P. falciparum*, and *P. vivax* among Humans and Anopheles dirus Mosquitoes, Southern Vietnam - Volume 17, Number 7—July 2011 - Emerging Infectious Diseases journal - CDC. Emerg Infect Dis. 2011;17(7):1232–9.

